# Association of Obstructive Sleep Apnea with Post-Acute Sequelae of SARS-CoV-2 infection (PASC)

**DOI:** 10.1101/2023.12.30.23300666

**Authors:** Stuart F. Quan, Matthew D. Weaver, Mark É. Czeisler, Laura K. Barger, Lauren A. Booker, Mark E. Howard, Melinda L. Jackson, Rashon I. Lane, Christine F. McDonald, Anna Ridgers, Rebecca Robbins, Prerna Varma, Joshua F. Wiley, Shantha M.W. Rajaratnam, Charles A. Czeisler

## Abstract

**Background:** Obstructive sleep apnea (OSA) is associated with COVID-19 infection. Fewer investigations have assessed OSA as a possible risk for the development of Post-Acute Sequelae of SARS-CoV-2 infection (PASC).

**Research Question:** In a general population, is OSA associated with increased odds of PASC-related symptoms and with an overall definition of PASC?

**Study Design:** Cross-sectional survey of a general population of 24,803 U.S. adults.

**Results:** COVID-19 infection occurred in 10,324 (41.6%) participants. Prevalence rates for a wide variety of persistent (> 3 months post infection) putative PASC-related physical and mental health symptoms ranged from 6.5% (peripheral edema) to 19.6% (nervous/anxious). In logistic regression models adjusted for demographic, anthropometric, comorbid medical and socioeconomic factors, OSA was associated with all putative PASC-related symptoms with the highest adjusted odds ratios (aOR) being fever (2.053) and nervous/anxious (1.939) respectively. Elastic net regression identified the 13 of 37 symptoms most strongly associated with COVID-19 infection. Four definitions of PASC were developed using these symptoms either weighted equally or proportionally by their regression coefficients. In all 4 logistic regression models using these definitions, OSA was associated with PASC (range of aORs: 1.934-2.071); this association was mitigated in those with treated OSA. In the best fitting overall model requiring ≥3 symptoms, PASC prevalence was 21.9%.

**Conclusion:** In a general population sample, OSA is associated with the development of PASC-related symptoms and a global definition of PASC. A PASC definition requiring the presence of 3 or more symptoms may be useful in identifying cases and for future research.

## Introduction

One devastating consequence of the COVID-19 pandemic has been the emergence of persistent and/or relapsing physical, cognitive and mental health symptoms beyond the acute phase of the illness commonly known as “long COVID” or more formally as Post-Acute Sequelae of SARS-CoV-2 infection (PASC).^1–4^ Prevalence estimates of PASC range from 7.5 to 41% in non-hospitalized adults depending on the criteria used to identify cases.^5^ The etiology of PASC has not been established although risk factors have been identified including greater severity of initial COVID-19 infection, multiple infections, female sex and preexisting health conditions.^4^

Obstructive sleep apnea (OSA) is a common sleep disorder characterized by repeated episodes of partial or complete obstruction of the upper airway during sleep. Recently, a number of studies have confirmed a strong association between OSA and COVID-19 infection.^6–11^ Intermittent hypoxia and sleep fragmentation resulting from upper airway obstruction can produce a milieu of chronic inflammation, oxidative stress, and immune system dysregulation,^12^ all of which have been implicated as mechanistic risk factors for developing COVID-19 infection;^13^ they may be important in the pathogenesis of PASC as well.^4^

Multiple studies have reported a high prevalence of symptoms observed with OSA among individuals with PASC, such as fatigue, insomnia and cognitive difficulties.^1,3,14^ Moreover, in a small prospective study of 60 COVID-19 survivors, OSA confirmed by a home sleep study was noted in 37 and was associated with a higher rate of abnormal pulmonary function and cognitive impairment.^15^ More recently, a retrospective analysis of data from three large health networks found an elevated risk of PASC in those with preexisting OSA.^16^ However, the association of OSA with specific symptoms associated with PASC was not explored. Nevertheless, these findings suggest that OSA could be a potential risk factor for the development and persistence of PASC symptoms.

Understanding the relationship between OSA and PASC is potentially important. It can help identify individuals who may be at a higher risk of developing PASC and may require closer monitoring and targeted interventions. Furthermore, it may provide insights into the underlying mechanisms that contribute to the persistence of COVID-19 symptoms thus informing the development of effective treatment strategies.^17^

In this study, we aimed to determine in a large general population cohort whether preexisting OSA is associated with the development of individual symptoms of PASC as well as with an overall definition of PASC based on frequency of symptoms. To accomplish this, we used data from the first five 2022 waves of The COVID-19 Outbreak Public Evaluation (COPE) Initiative (http://www.thecopeinitiative.org/), a program focused on accumulating data on public attitudes, behaviors and beliefs related to the COVID-19 pandemic from large scale, demographically representative samples.

## Methods

### Study Design and Participants

From March 10, 2022 to October 15, 2022, the COPE Initiative administered five successive cross-sectional waves surveys focused on accumulating data on the prevalence and sequelae of COVID-19 infection. Dates of administration were Wave 1 (March 10-30, 2022), Wave 2 (April 4-May 1, 2022), Wave 3 (May 4-June 2, 2022), Wave 4 (August 1-18, 2022), Wave 5 (September 26-October 15, 2022); each wave consisted of more than 5000 unique participants who were recruited to approximate population estimates for age, sex, race, and ethnicity based on the 2020 U.S. census. Surveys were conducted online by Qualtrics, LLC (Provo, Utah, and Seattle, Washington, U.S.), using their network of participant pools with varying recruitment methodologies that include digital advertisements and promotions, word-of-mouth and membership referrals, social networks, television and radio advertisements, and offline mail-based approaches. Informed consent was obtained electronically. The study was approved by the Monash University Human Research Ethics Committee (Study #24036).

### Survey Items

Participants self-reported demographic, anthropometric, and socioeconomic information including age, race, ethnicity, sex, height and weight, education level, employment status and household income. In addition, they reported information on several current and past medical conditions by answering the question: “Have you ever been diagnosed with any of the following conditions?” In addition to OSA, opportunity was provided to endorse high blood pressure, cardiovascular disease (e.g., heart attack, stroke, angina), gastrointestinal disorder (e.g., acid reflux, ulcers, indigestion), cancer, chronic kidney disease, liver disease, sickle cell disease, chronic obstructive pulmonary disease, and asthma. Possible responses to each condition were “Never”, “Yes I have in the past, but don’t have it now”, “Yes I have, but I do not regularly take medications or receiving treatment”, and “Yes I have, and I am regularly taking medications or receiving treatment”.

COVID-19 vaccination status was ascertained by asking “How many COVID-19 vaccine doses have you received? (If you have had two doses of one brand and one of another, please select three)”. Participants were allowed to respond from 0 to 4.

Symptoms of OSA were obtained from responses to the Pittsburgh Sleep Quality Index which was embedded into each survey and included items related to roommate or bedpartner reported “loud snoring” and “long pauses between breaths while you sleep”.^18^ In addition, sleepiness was assessed from the following item in the questionnaire: During the past month, how often have you had trouble staying awake while driving, eating meals, or engaging in social activity”. Possible responses to all three items were “Not during the past month”, “less than once a week”, Once or twice a week” or “Three or more times a week”. Participants were considered to have symptoms of OSA if they had either of the following combination of symptoms: 1) snoring “Three or more times a week” and witnessed apnea or sleepiness “Once or twice a week”; 2) witnessed apnea and sleepiness “Once or twice a week”.

Each survey contained identical items related to COVID-19 infection status and the number of COVID-19 vaccinations participants had obtained. Ascertainment of past COVID-19 infection was obtained using responses from following questions related to COVID-19 testing or the presence of loss of taste or smell:

1. “Have you ever tested positive?”
2. “Despite never testing positive, are you confident that you have had COVID-19?”
3. “Despite never testing positive, have you received a clinical diagnosis of COVID- 19?”
4. “Have you experienced a problem with decreased sense of smell or taste at any point since January 2020?”

Identical items pertaining to symptoms potentially associated with PASC were included in each survey. Participants who endorsed having had any of aforementioned items #1-4 were asked to provide the date of their positive test or onset of infection. Additionally, they were asked if they experienced any of the following general health symptoms more than 2 weeks after their infection: fever, sweats or chills; tired/fatigued; loss of smell (anosmia); loss of taste (dysgeusia); runny nose or congestion; sore throat; ringing or other noises in the ear (tinnitus); shortness of breath (dyspnea); cough or sputum production; nausea; uneasiness or discomfort; diarrhea or constipation; chest pain; heart palpitations; peripheral edema (swelling of lower legs or hands); headache; excessive sleepiness; problems sleeping; limited physical activity or exercise; limited social activity (such as meeting friends or family); other aches or pains. For each symptom, participants who endorsed having experienced the symptom more than 2 weeks after their COVID-19 infection or indicated that they were currently having the symptom were asked how long their symptoms persisted after their infection. Response options were less than one month, 1-2 months, 3-6 months, 6-12 months, greater than 12 months.

Similarly, participants who endorsed having had a positive COVID-19 test or infection were asked if they experienced any of the following cognitive and mental health symptoms more than 2 weeks after their infection: forgetful; difficulty thinking; difficulty focusing; cloudy; difficulty finding the right words/communicating; mental fatigue; slow; mind went blank; feeling nervous or anxious; feeling agitated; believing people are trying to harm you; unable to control worrying; little pleasure in doing things; feeling depressed; sleep difficulties; irritability; and avoiding places, people or situations that are reminders of COVID-19 disease. The duration of each symptom with a positive response was also solicited utilizing response options identical to those used for general health symptoms.

In parallel to participants presumed to have had COVID-19, those who never had a positive test for COVID-19, who affirm never having had a COVID-19 infection, and who never had a loss of taste or smell were asked whether they experienced any of the same general health, cognitive or mental health symptoms in the two weeks preceding the survey. However, they were not queried regarding duration of any positive symptoms.

### Statistical Analyses

As in our previously analyses in this cohort, we defined a positive history of COVID-19 infection as an affirmative response to having tested positive for COVID-19, a clinical diagnosis of COVID-19, or loss of taste or smell.^11,19^ Participants were considered to have OSA if they endorsed currently having the condition whether treated or not or if they had two or more symptoms of OSA.^11^ Vaccination status was dichotomized as Boosted (>2 vaccinations) or Not Boosted ( ≤ 2 vaccinations).^11^ Comorbid medical conditions were defined as currently having the condition whether treated or untreated. The effect of comorbid medical conditions was evaluated by summing the number of conditions reported by the participant (minimum value 0, maximum value 9).^11^ Body mass index (BMI) was calculated using self-reported height and weight as kg/m^2^. Socioeconomic covariates were dichotomized as follows: employment (retired vs. not retired), education (high school or less vs. some college) and annual income in U.S. Dollars (<$50,000 vs ≥$50,000). *Participants were considered to have a general health, cognitive or mental health symptom potentially associated with PASC if they endorsed having the symptom for at least 3 months after their COVID-19 infection*.

Summary data for continuous or ordinal variables are reported as their respective means and standard deviations (SD) and for categorical variables as their percentages. Comparisons of co-morbid medical, demographic, and social characteristic variables stratified by COVID-19 infection status were performed using Student’s unpaired t-test for continuous or ordinal variables and χ^2^ for categorical variables. For individual symptoms potentially associated with PASC, a z test for proportions was used to assess differences among COVID-19 infection groups.

Multivariable modelling using logistic regression was utilized to determine whether OSA was associated with each individual symptom potentially associated with PASC among COVID-19 positive participants. For each symptom, a baseline model was constructed using only OSA. We then developed increasingly complex models by sequentially including demographic factors, comorbidities, boosted vaccination status, and socioeconomic factors. Results of the logistic regression models are presented as unadjusted or adjusted odds ratios (aOR) and their 95% confidence intervals (95% CI).

To assess whether OSA was associated with PASC overall rather than individual symptoms, we developed several different models of PASC based on symptom frequency. Initially, using positive COVID-19 status as a binary outcome, we performed an elastic net regression using 10-fold cross-validation to mitigate the impact of multicollinearity and select the most relevant symptoms that would contribute to an overall model of PASC. After fitting the elastic net regression model, we obtained regression coefficients for each predictor symptom. Symptoms with non-zero coefficients were considered the most important predictors associated with PASC, while symptoms with zero coefficients were deemed as less influential or irrelevant and were eliminated from further modelling considerations. Next, a PASC score was calculated using a multiple linear regression using the symptom variables selected and weighted by the elastic net procedure. For two models, overall PASC (symptoms present > 3 months) was defined as a score exceeding the 95^th^ and 99^th^ percentiles respectively of the calculated scores of participants who had no evidence for COVID-19 infection. For two additional models, the symptom variables were summed (maximum value 13); overall PASC was defined as a score of ≥2 and ≥3 which represent the 97.5th and 99th percentiles respectively of scores of participants who had no evidence for COVID-19 infection. Finally, logistic regression was performed in the group of participants with evidence of COVID-19 infection for each of the 4 PASC models to determine the association between OSA and overall PASC after adjusting for boosted vaccination status, demographic factors, comorbidities, and socioeconomic factors.

To determine whether our definition of COVID-19 infection status influenced our results, we performed sensitivity analyses with stricter (i.e., using COVID-19 infection as a positive test only) and broader (i.e., our original definition plus assumed positive for COVID-19 without a positive test as an indicator of a past COVID-19 infection). Additional sensitivity analyses were conducted with an OSA definition that omitted participants with OSA symptoms but did not self-report of a diagnosis of OSA.

All analyses were conducted using IBM SPSS version 28 (Armonk, NY). A p<0.05 was considered statistically significant.

## Results

Table 1 shows the associations between COVID-19 infection status and various co-morbid medical conditions, demographic, and social characteristics of the 24,803 participants in the cohort. Groups who had experienced COVID-19 infection (N=10,324, 41.6%) were younger age, less commonly retired and had greater income; Hispanics were at greater risk than other racial or ethnic groups. They also had a greater number of comorbidities and had a higher prevalence of OSA.

The prevalence rates of symptoms putatively associated with PASC are presented in Table 2. Participants without any evidence of having been infected with COVID-19 had very low prevalence rates of all symptoms particularly in comparison to participants who had previously experienced COVID-19 infection (all p values <0.0001). In addition, the prevalence rates of COVID-19 negative participants were also lower than those participants who had symptoms persisting for greater than 3 months (all p values <0.0001). Among COVID-19 participants with persistent symptoms, fatigue (15.2%) and loss of smell (12.5%) and taste (11.4%) had the highest prevalence rates among general health symptoms; peripheral edema (6.5%) had the lowest prevalence. However, even more common was the occurrence of mental health symptoms with the prevalence of nervousness or anxiety approaching 1 in 5 COVID-19 infected participants.

Logistic regression models of the association between OSA and individual symptoms putatively associated with PASC are displayed in Table 3. Obstructive sleep apnea was associated with a greater likelihood of all of symptoms in unadjusted, partially, and fully adjusted models. The highest aORs for general health, cognitive and mental health symptoms were fever (2.053), mind went blank (1.669) and feeling nervous/anxious (1.939) respectively.

In Table 4 is shown the standardized elastic net regression coefficients for symptoms possibly associated with PASC. Of the initial 37 symptoms entered into the elastic net regression, only 13 had non-zero coefficients. Fever, fatigue, and loss of smell or taste had the highest coefficients. With the exception of forgetful, the remaining cognitive and mental health symptoms had relatively small contributions to the overall regression.

Table 5 presents the logistic regression models of the association between OSA and four models of PASC as constructed from the results of the elastic net regression. The number of participants and PASC prevalence rates for each model are provided as well. Prevalence rates ranged from 21.8% (at least 2 symptoms present) to 38.5% (95^th^ percentile of PASC scores). For all four models, unadjusted, partially, and fully adjusted aORs demonstrated that OSA was associated with a higher likelihood of having PASC. The aOR for the best fitting model (≥3 symptoms present) was 2.059 (95% CI: 1.769- 2.395). The remaining models had similar aORs. In addition, boosted vaccination status was protective against development of PASC (aOR for ≥3 symptoms present: 0.776, 95% CI: 0.656-0.918).

In sensitivity analyses (Table S1) both a definition of COVID-19 that was stricter (positive test, loss of taste or smell) and more liberal (positive test, loss of taste or smell, clinical diagnosis but no positive test) demonstrated a strong association of OSA with all four models of PASC. Current or past treatment of OSA also showed a strong relationship with PASC. However, current treatment of OSA alone was not associated with PASC in all four models.

## Discussion

Our analyses found that the prevalence of symptoms heretofore associated with PASC was much higher than in individuals who never had a COVID-19 infection. Furthermore, all of these symptoms individually were associated with OSA after adjustment for demographic and socioeconomic covariates and medical comorbidities. In several models of PASC utilizing definitions based on number of symptoms experienced, OSA was consistently associated with PASC. Further analyses indicated that this association was mitigated by OSA treatment. Additionally, boosted vaccination was protective against the development of PASC.

A large number of symptoms involving nearly every organ system are reported by individuals who fail to completely recover many weeks after a COVID-19 infection and who are thus diagnosed as having PASC.^1,3,20^ For many of these commonly reported physical and mental health symptoms, we now document that they are highly prevalent more than 3 months afterwards among individuals who previously experienced a COVID-19 infection in comparison to those who have never been infected. Consistent with recent reviews, we found that fatigue and dysgeusia or dysosmia were among the most frequently reported symptoms among those with PASC.^1,3^ In contrast, persistent fever was a common occurrence among our participants whereas it was less prominent in other studies.^3^ In an extension to other reports, our findings also highlight the variety of persistent mental health symptoms after a COVID-19 infection. The high rates of depression, anxiety and generalized dysphoria are likely factors that explain a large portion of the poor quality of life experienced by individuals with PASC.^21^

We observed that OSA was associated with all of the symptoms linked with PASC in our study after controlling for demographic, medical comorbidities, and variation in socioeconomic factors. We and others have documented that OSA appears to be a risk factor for COVID-19 infection.^6,7,9–11^ In a recent study using an amalgam of three different definitions of PASC identified from electronic medical records, an association with OSA was found with an adjusted odds ratio of 1.75.^16^ Our findings extend this observation by documenting the association of OSA with individual symptoms of PASC. In contrast to the prevalence rates of other PASC symptoms particular those associated with mental health, fever had the strongest relationship with OSA whereas it was somewhat weaker for fatigue. This latter observation may be explained by the common occurrence of fatigue as a symptom of OSA.

Our elastic net regression results identified 13 symptoms that were strongly related to a diagnosis of COVID-19. This demonstrates that there is a substantial amount of overlap in the strength of various PASC related symptoms and their association with a diagnosis of COVID-19. In contradistinction to our findings that mental health symptoms were highly prevalent among participants more than 3 months after their infection, the physical symptoms of fever, fatigue, dysgeusia and dysosmia had the strongest association with infection. In a recent study from the RECOVER cohort in which a lasso regression was used for selection of the most important PASC related symptoms, abnormal smell or taste also was the strongest predictor.^22^ Thus, our results are consistent with this prior study, but differ in finding that fatigue and fever also had a high predictive value. The differences may relate to the composition of the cohorts; the COPE initiative cohort was recruited from the general population whereas RECOVER primarily targeted individuals who had been infected with COVID-19.

Although there is general consensus that PASC consists of a constellation of symptoms, there is no uniform definition of the syndrome or diagnostic biomarker.^14^ Recently, a definition of PASC was proposed using data from the RECOVER cohort in which points were allocated among 12 symptoms derived after applying a lasso regression to 37 candidate symptoms. Dysguesia/Dysosmia and post exertional malaise were allocated the most points, 8 and 7 respectively, whereas fatigue was allocated only 1 point. A score of 12 or greater was considered diagnostic of PASC; using this definition, the prevalence of PASC was 23%. In contrast, using a different methodology, we found a similar prevalence rate using our two models that employed a more stringent definition, 97.5^th^ percentile (21.8%) and at least 3 positive symptoms (21.9%) (Table 5). We propose that the latter definition will be easier to implement than the more complex algorithm derived from the RECOVER cohort.^22^ However, further studies to validate this approach will be necessary.

Obstructive sleep apnea was found to be associated with PASC in all 4 of the models that we developed. Thus, our results replicate the observations from the RECOVER cohort in which they also noted a strong association between PASC and OSA using an approach that identified participants with PASC based on medical record documentation.^16^ We extend this finding by demonstrating that there was no association between OSA and PASC in participants who had been treated for their OSA. This is consistent with our previous observation that risk of COVID-19 infection was reduced in those with treated OSA.^11^

We observed that boosted vaccination status was protective against the development of PASC in our models. This finding is consistent with the conclusion from a recent systematic review of the protective effects of COVID-19 infection.^23^ It provides additional support for public health messaging concerning the importance of COVID-19 infection.

The mechanism underlying the association between OSA and PASC remains to be determined. However, there are several possibilities. Substantial evidence supports that OSA is an inflammatory condition.^12^ Intermittent hypoxemia resulting from OSA promotes the release of inflammatory cytokines such as interleukin-6, c-reactive protein, and tumor necrosis factor-α.^24^ Introduction of the SARS-CoV-2 virus into a pre-existing inflammatory milieu could increase the likelihood of more severe COVID-19 outcomes; severe COVID-19 is associated with an increased risk of PASC.^4^ There also is evidence that intermittent hypoxemia is responsible for the existence of a hypercoagulable state in OSA.^25,26^ Microthrombi have been observed in PASC and have been implicated as contributing to both neurologic and cardiovascular symptoms.^27^ Intermittent hypoxemia also produces oxidative stress with release of reactive oxygen species.^28^ A hypercoagulable state, inflammation and oxidative stress are all factors that individually or in combination can lead to disruption of the blood brain barrier resulting in the cognitive and neurologic symptoms of PASC.^14,26^ In addition, abnormalities in cellular immunity have been documented in PASC;^14^ OSA also has been associated with cellular immune dysfunction and therefore may be a contributing factor to the abnormal immune function in PASC.^29^

We acknowledge that our results are subject to some limitations. Identification of COVID-19 and subsequent symptoms as well as OSA were self-reported which may have resulted in misclassification of the exposure and/or outcome. Although it is possible that OSA was reported more often in persons who had experienced a COVID-19 infection, we believe this is unlikely inasmuch as this linkage is not well known among the general public. Furthermore, in our sensitivity analyses, changes in the definition of COVID-19 resulted in similar findings. We also acknowledge that in our fully adjusted models, residual confounding may have been present. However, we adjusted for multiple comorbid medical conditions including obesity (via BMI) and diabetes; the latter are well-accepted risk factors for COVID-19 infection. Finally, with the exception of the recent study from the RECOVER cohort,^22^ there have been no other definitions of PASC. Thus, the definitions utilized in our modeling of PASC were to some extent arbitrary. However, the use of percentiles ranging from the 95^th^ to 99^th^ is a common practice to define normality in anthropometry and other health conditions such as obesity.^30^ In contrast to these limitations, our study has two major strengths, the size of cohort (N=24,803) and that it was generally representative of the U.S. adult population.

In conclusion, there is a strong association between OSA and symptoms experienced by individuals more than 3 months after a COVID-19 infection. In several model definitions of PASC, OSA was strongly predictive after adjustment for demographic, anthropometric, comorbid medical conditions, and socioeconomic factors. This relationship was mitigated with OSA treatment. Therefore, OSA should be considered an important risk factor for the development of PASC. Furthermore, the presence of 3 or more PASC related symptoms is a practical definition of PASC, but additional validation is required.

## Data Availability

All data produced in the present study are available upon reasonable request to the authors.

## Acknowledgments

Concept and Design: SFQ

Data collection: MDW, MÉC, MEH

Data analysis and interpretation: SFQ, MDW, MÉC, LAB, MEH

Drafting of the manuscript: SFQ

Critical feedback and revision of manuscript: SFQ, MDW, MÉC, LKB, LAB, MEH, MLJ, RL, CFM, AR, RR, PV, SMWR, CAC

