## Supplementary material for "Association of Obstructive Sleep Apnea with Post-Acute Sequelae of SARS-CoV-2 infection (PASC)": Table 1

Table 1: Associations Between COVID-19 Infection Status and Obstructive Sleep Apnea,

Co-morbid Medical, Demographic and Social Characteristics^*^

|  | | **COVID-19 Negative**  **(N=14479)** | | **COVID-19**  **Positive**  **(N=10324)** | | **COVID-19**  **Overall**  **(N=24803)** | |
| --- | --- | --- | --- | --- | --- | --- | --- |
|  | | *Mean* | *SD* | *Mean* | *SD* | *Mean* | *SD* |
| Age (y) ^*^ | | 50.6 | 18.0 | 39.3 | 15.3 | 46.6 | 17.8 |
| Body Mass Index (kg/m^2^) | | 27.5 | 5.9 | 27.6 | 6.3 | 28.4 | 8.9 |
| No. Comorbidities ^*^ | | 1.1 | 1.5 | 2.1 | 2.9 | 1.1 | 1.8 |
|  | | *N* | *%* | *N* | *%* | *N* | *%* |
| Sex | |  |  |  |  |  |  |
|  | Male | 7027 | 48.8 | 5007 | 49.1 | 12034 | 48.9 |
|  | Female | 7385 | 51.2 | 5198 | 50.9 | 12583 | 51.1 |
| Race/Ethnicity ^*^ | |  |  |  |  |  |  |
|  | White | 9206 | 63.6 | 6032 | 58.4 | 15238 | 61.4 |
|  | Black | 1581 | 10.9 | 1056 | 10.2 | 2637 | 10.6 |
|  | Asian | 1024 | 7.1 | 513 | 5.0 | 1537 | 6.2 |
|  | Hispanic | 1926 | 13.3 | 2264 | 21.9 | 4190 | 16.9 |
|  | Other | 742 | 5.1 | 459 | 4.4 | 1201 | 4.8 |
| Employment ^*^ | |  |  |  |  |  |  |
|  | Retired | 4471 | 30.9 | 1382 | 13.4 | 5853 | 23.6 |
|  | Not Retired | 10008 | 69.1 | 8942 | 86.6 | 18950 | 76.4 |
| Education | |  |  |  |  |  |  |
|  | High School or Less | 3864 | 26.7 | 2670 | 25.9 | 6534 | 26.3 |
|  | Some College | 10615 | 73.3 | 7654 | 74.1 | 18269 | 73.7 |
| Income (Yearly) ^*^ | |  |  |  |  |  |  |
|  | < $50,000 | 6621 | 48.1 | 4136 | 41.3 | 10757 | 45.2 |
|  | ≥ $50,000 | 7156 | 51.9 | 5873 | 58.7 | 13029 | 54.8 |
| Vaccination Boosted | |  |  |  |  |  |  |
|  | No (≤2 Vaccinations) | 7726 | 55.5 | 7207 | 71.9 | 14933 | 62.3 |
|  | Yes (>2 Vaccinations) | 6198 | 44.5 | 2823 | 28.1 | 9021 | 37.7 |
| Obstructive Sleep Apnea^*^ | |  |  |  |  |  |  |
|  | Yes | 1756 | 12.1 | 2828 | 27.4 | 4584 | 18.5 |
|  | No | 12723 | 87.9 | 7496 | 72.6 | 20219 | 81.5 |

^*^Minor discrepancies in case counts and totals reflect small amounts of missing data and rounding

Significant differences in means or proportions: ^*^p≤0.001
