## Supplementary material for "Association of Obstructive Sleep Apnea with Post-Acute Sequelae of SARS-CoV-2 infection (PASC)": Table 2

**Table 2: Prevalence of Candidate Symptoms Associated with PASC Stratified by COVID-19 Infection Status**

| | COVID-19 Negative N=14479 | | COVID-19 Positive N=10324 | | COVID-19 Symptoms > 3 months | |
|---|---|---|---|---|---|---|
| | *N* | *%* | *N* | *%* | *N* | *%* |
| *GENERAL HEALTH SYMPTOMS** | | | | | | |
| Fever | 77 | 0.5 | 5339 | 51.7 | 1110 | 10.8 |
| Fatigue | 451 | 3.1 | 5571 | 54.0 | 1569 | 15.2 |
| Loss of Smell | 28 | 0.2 | 4623 | 44.8 | 1291 | 12.5 |
| Loss of Taste | 21 | 0.1 | 4502 | 43.6 | 1177 | 11.4 |
| Nasal Congestion | 386 | 2.7 | 4693 | 45.5 | 1006 | 9.7 |
| Sore Throat | 151 | 1.0 | 4472 | 43.3 | 866 | 8.4 |
| Tinnitus | 222 | 1.5 | 3619 | 35.1 | 936 | 9.1 |
| Dyspnea | 116 | 0.8 | 3989 | 38.6 | 1089 | 10.5 |
| Cough/Sputum Production | 183 | 1.3 | 4233 | 41.0 | 957 | 9.3 |
| Nausea | 133 | 0.9 | 3658 | 35.4 | 813 | 7.9 |
| Uneasieness/Discomfort | 89 | 0.6 | 3756 | 36.4 | 980 | 9.5 |
| Diarrhea/Constipation | 182 | 1.3 | 3467 | 33.6 | 782 | 7.6 |
| Chest Pain | 63 | 0.4 | 3296 | 31.9 | 796 | 7.7 |
| Palpitations | 47 | 0.3 | 3074 | 29.8 | 776 | 7.5 |
| Peripheral Edema | 56 | 0.4 | 2855 | 27.7 | 675 | 6.5 |
| Headache | 410 | 2.8 | 4084 | 39.6 | 993 | 9.6 |
| Sleepiness | 311 | 2.1 | 3632 | 35.2 | 675 | 6.5 |
| Sleep Problems | 315 | 2.2 | 3436 | 33.3 | 1034 | 10.0 |
| Limited Physical Activity | 189 | 1.3 | 3707 | 35.9 | 1073 | 10.4 |
| Limited Social Activity | 182 | 1.3 | 3669 | 35.5 | 969 | 9.4 |
| Other Aches or Pains | 258 | 1.8 | 3418 | 33.1 | 973 | 9.4 |
| *COGNITIVE SYMPTOMS** | | | | | | |
| Forgetful | 360 | 2.5 | 4205 | 40.7 | 1185 | 11.5 |
| Difficulty Thinking | 199 | 1.4 | 3989 | 38.6 | 1224 | 11.9 |
| Difficulty Focusing | 274 | 1.9 | 3911 | 37.9 | 1165 | 11.3 |
| Cloudy | 155 | 1.1 | 3749 | 36.3 | 1075 | 10.4 |
| Difficulty Finding Words | 245 | 1.7 | 3526 | 34.2 | 1028 | 10.0 |
| Mental Fatigue | 322 | 2.2 | 3783 | 36.6 | 1153 | 11.2 |
| Slow | 139 | 1.0 | 3481 | 33.7 | 1012 | 9.8 |
| Mind Went Blank | 269 | 1.9 | 3388 | 32.8 | 1078 | 10.4 |
| *MENTAL HEALTH SYMPTOMS** | | | | | | |
| Feeling Nervous/Anxious | 735 | 5.1 | 3536 | 34.3 | 2021 | 19.6 |
| Feeling Agitated | 477 | 3.3 | 2431 | 23.5 | 1904 | 18.4 |
| Being Harmed by People | 88 | 0.6 | 2287 | 22.2 | 1265 | 12.3 |
| Inability to Control Worrying | 334 | 2.3 | 2148 | 20.8 | 1716 | 16.6 |
| Little Pleasure | 403 | 2.8 | 2472 | 23.9 | 1750 | 17.0 |
| Depressed | 601 | 4.2 | 2459 | 23.8 | 1915 | 18.5 |
| Irritability | 401 | 2.8 | 2230 | 21.6 | 1701 | 16.5 |
| Avoiding People/Places | 185 | 1.3 | 2021 | 19.6 | 1605 | 15.5 |

*All symptoms higher in COVID-19 Positive and COVID-19 Symptoms >3 months Groups
