## Supplementary material for "Association of Obstructive Sleep Apnea with Post-Acute Sequelae of SARS-CoV-2 infection (PASC)": Table 3

**Table 3: Logistic Regression Models of the Association Between Obstructive Sleep Apnea and Individual Candidate Symptoms of PASC**

| | Unadjusted | | | Model 1 | | | Model 2 | | | Model 3 | | |
|---|---|---|---|---|---|---|---|---|---|---|---|---|
| | *OR* | *95% CI* | | *aOR* | *95% CI* | | *aOR* | *95% CI* | | *aOR* | *95% CI* | |
| | | *Lower* | *Upper* | | *Lower* | *Upper* | | *Lower* | *Upper* | | *Lower* | *Upper* |
| *GENERAL HEALTH SYMPTOMS** | | | | | | | | | | | | |
| Fever | 4.429 | 3.895 | 5.037 | 4.083 | 3.579 | 4.659 | 2.087 | 1.714 | 2.541 | 2.053 | 1.682 | 2.505 |
| Fatigue | 2.509 | 2.245 | 2.803 | 2.544 | 2.271 | 2.850 | 1.504 | 1.269 | 1.783 | 1.494 | 1.259 | 1.774 |
| Loss of Smell | 3.203 | 2.843 | 3.609 | 3.052 | 2.701 | 3.449 | 1.778 | 1.484 | 2.131 | 1.780 | 1.483 | 2.138 |
| Loss of Taste | 2.928 | 2.634 | 3.374 | 2.932 | 2.582 | 3.329 | 1.921 | 1.594 | 2.315 | 1.928 | 1.598 | 2.328 |
| Nasal Congestion | 3.512 | 3.076 | 4.010 | 3.303 | 2.892 | 3.794 | 1.640 | 1.337 | 2.012 | 1.607 | 1.308 | 1.975 |
| Sore Throat | 4.206 | 3.647 | 4.852 | 3.831 | 3.309 | 4.434 | 1.674 | 1.338 | 2.094 | 1.642 | 1.311 | 2.057 |
| Tinnitus | 3.394 | 2.961 | 3.891 | 3.177 | 2.862 | 3.654 | 1.552 | 1.258 | 1.916 | 1.559 | 1.260 | 1.929 |
| Dyspnea† | 2.862 | 2.519 | 3.252 | 2.802 | 2.458 | 3.195 | 1.371 | 1.122 | 1.676 | 1.363 | 1.114 | 1.668 |
| Cough/Sputum Production† | 3.308 | 2.890 | 3.787 | 3.130 | 2.724 | 3.597 | 1.382 | 1.120 | 1.706 | 1.383 | 1.119 | 1.709 |
| Nausea | 3.698 | 3.196 | 4.278 | 3.462 | 2.979 | 4.024 | 1.906 | 1.520 | 2.389 | 1.913 | 1.523 | 2.403 |
| Uneasieness/Discomfort | 2.808 | 2.456 | 3.209 | 2.711 | 2.363 | 3.110 | 1.440 | 1.171 | 1.772 | 1.442 | 1.170 | 1.776 |
| Diarrhea/Constipation | 3.921 | 3.379 | 4.551 | 3.703 | 3.178 | 4.314 | 1.778 | 1.415 | 2.234 | 1.775 | 1.410 | 2.235 |
| Chest Pain | 3.474 | 2.999 | 4.023 | 3.260 | 2.804 | 3.791 | 1.582 | 1.265 | 1.978 | 1.586 | 1.266 | 1.986 |
| Palpitations | 3.926 | 3.382 | 4.559 | 3.727 | 3.198 | 4.344 | 1.664 | 1.320 | 2.096 | 1.651 | 1.307 | 2.084 |
| Peripheral Edema | 4.184 | 3.566 | 4.909 | 3.906 | 3.315 | 4.603 | 1.623 | 1.265 | 2.083 | 1.621 | 1.260 | 2.085 |
| Headache | 2.503 | 2.191 | 2.859 | 2.463 | 2.148 | 2.824 | 1.514 | 1.234 | 1.858 | 1.517 | 1.235 | 1.865 |
| Sleepiness | 2.583 | 2.206 | 3.025 | 2.479 | 2.108 | 2.916 | 1.490 | 1.169 | 1.898 | 1.520 | 1.191 | 1.940 |
| Sleep Problems | 2.461 | 2.159 | 2.805 | 2.446 | 2.138 | 2.797 | 1.345 | 1.100 | 1.646 | 1.367 | 1.116 | 1.675 |
| Limited Physical Activity | 2.464 | 2.166 | 2.803 | 2.481 | 2.173 | 2.831 | 1.421 | 1.164 | 1.735 | 1.449 | 1.185 | 1.772 |
| Limited Social Activity‡ | 2.415 | 2.111 | 2.764 | 2.397 | 2.087 | 2.752 | 1.273 | 1.033 | 1.571 | 1.289 | 1.043 | 1.591 |
| Other Aches or Pains | 2.523 | 2.206 | 2.886 | 2.529 | 2.020 | 2.904 | 1.586 | 1.291 | 1.949 | 1.605 | 1.304 | 1.974 |
| *COGNITIVE SYMPTOMS** | | | | | | | | | | | | |
| Forgetful | 2.448 | 2.163 | 2.770 | 2.594 | 2.284 | 2.946 | 1.596 | 1.321 | 1.928 | 1.625 | 1.343 | 1.965 |
| Difficulty Thinking | 2.641 | 2.338 | 2.983 | 2.728 | 2.407 | 3.092 | 1.527 | 1.266 | 1.842 | 1.534 | 1.269 | 1.853 |
| Difficulty Focusing | 2.206 | 1.947 | 2.500 | 2.324 | 2.043 | 2.644 | 1.380 | 1.137 | 1.675 | 1.375 | 1.131 | 1.673 |
| Cloudy | 2.465 | 2.167 | 2.804 | 2.544 | 2.230 | 2.902 | 1.560 | 1.283 | 1.898 | 1.588 | 1.303 | 1.936 |
| Difficulty Finding Words | 2.126 | 1.863 | 2.426 | 2.239 | 1.955 | 2.565 | 1.425 | 1.165 | 1.742 | 1.439 | 1.175 | 1.763 |
| Mental Fatigue | 2.081 | 1.835 | 2.361 | 2.253 | 1.978 | 2.565 | 1.401 | 1.154 | 1.700 | 1.426 | 1.173 | 1.733 |
| Slow | 2.557 | 2.241 | 2.918 | 2.618 | 2.287 | 2.998 | 1.553 | 1.267 | 1.903 | 1.574 | 1.282 | 1.932 |
| Mind Went Blank | 2.431 | 2.137 | 2.765 | 2.525 | 2.213 | 2.883 | 1.666 | 1.370 | 2.026 | 1.669 | 1.370 | 2.033 |
| *MENTAL HEALTH SYMPTOMS** | | | | | | | | | | | | |
| Feeling Nervous/Anxious | 2.952 | 2.661 | 3.274 | 2.960 | 2.659 | 3.296 | 1.962 | 1.678 | 2.294 | 1.939 | 1.655 | 2.271 |
| Feeling Agitated | 3.247 | 2.920 | 3.611 | 3.220 | 2.886 | 3.593 | 1.838 | 1.563 | 2.160 | 1.800 | 1.528 | 2.120 |
| Being Harmed by People | 4.192 | 3.701 | 4.748 | 3.839 | 3.377 | 4.363 | 1.899 | 1.570 | 2.298 | 1.873 | 1.545 | 2.271 |
| Inability to Control Worrying | 2.858 | 2.560 | 3.189 | 2.829 | 2.527 | 3.168 | 1.680 | 1.421 | 1.986 | 1.668 | 1.409 | 1.975 |
| Little Pleasure | 2.740 | 2.457 | 3.056 | 2.756 | 2.463 | 3.083 | 1.516 | 1.282 | 1.793 | 1.520 | 1.284 | 1.800 |
| Depressed | 2.427 | 2.182 | 2.700 | 2.473 | 2.215 | 2.761 | 1.440 | 1.223 | 1.695 | 1.449 | 1.229 | 1.708 |
| Irritability | 2.453 | 2.196 | 2.740 | 2.484 | 2.216 | 2.784 | 1.429 | 1.207 | 1.692 | 1.440 | 1.215 | 1.708 |
| Avoiding People/Places | 2.715 | 2.426 | 3.038 | 2.712 | 2.415 | 3.046 | 1.557 | 1.309 | 1.852 | 1.560 | 1.309 | 1.858 |

Model 1: adjusted for age, sex and race
Model 2: Model 1 + bmi, comorbidities, boosted COVID-19 vaccination status, and boosted COVID-19 vaccination status/OSA interaction
Model 3: Model 2 + income, employment and education

*p <0.001 for association of each symptom and OSA except for dyspnea, cough, sleep problems and limited social activity
†p <0.01 for association of dyspnea, cough and sleep problems and OSA for Models 2 and 3
‡p <0.05 for association of limited social activity and OSA for Models 2 and 3
