## Supplementary material for "Association of Obstructive Sleep Apnea with Post-Acute Sequelae of SARS-CoV-2 infection (PASC)": Table 4

**Table 4: Elastic Net Regression Standardized Coefficients for Symptoms Associated with PASC**

| | Standardized Coefficient |
|---|---|
| Fever | 0.048 |
| Fatigue | 0.031 |
| Loss of Smell | 0.026 |
| Loss of Taste | 0.018 |
| Sore Throat | 0.001 |
| Sleepiness | 0.001 |
| Forgetful | 0.013 |
| Difficulty Thinking | 0.011 |
| Difficulty Focusing | 0.006 |
| Cloudy | 0.004 |
| Slow | 0.003 |
| Feeling Nervous/Anxious | 0.003 |
| Avoiding People/Places | 0.003 |
| Intercept | 0.416 |
| L1 Ratio | 0.1 |
| Alpha | 0.1 |
| Dependent Variable | Previous COVID-19 Infection |
