## Supplementary material for "Association of Obstructive Sleep Apnea with Post-Acute Sequelae of SARS-CoV-2 infection (PASC)": Table 5

**Table 5: Logistic Regression Models of the Association of Obstructive Sleep Apnea with Overall PASC Models**

| | **PASC Prevalence** | **Unadjusted** | | | **Model 1** | | | **Model 2** | | | **Model 3** | | | **Model 3 Goodness of Fit and Variance** | |
|---|---|---|---|---|---|---|---|---|---|---|---|---|---|---|---|
| | | *OR* | *95% CI* | | *aOR* | *95% CI* | | *aOR* | *95% CI* | | *aOR* | *95% CI* | | -2 Log likelihood | Nagelkerke R2 |
| **PASC Model*** | *N/%* | | *Lower* | *Upper* | | *Lower* | *Upper* | | *Lower* | *Upper* | | *Lower* | *Upper* | | |
| PASC: 97.5th Percentile | 2260/21.8 | 3.451 | 3.128 | 3.807 | 3.388 | 3.063 | 3.747 | 1.940 | 1.671 | 2.253 | 1.934 | 1.663 | 2.249 | 8078.772 | 0.140 |
| PASC: 95th Percentile | 3976/38.5 | 3.770 | 3.444 | 4.127 | 3.740 | 3.406 | 4.106 | 2.059 | 1.795 | 2.361 | 2.024 | 1.762 | 2.325 | 10067.928 | 0.185 |
| PASC: At Least 2 Positive | 2985/28.9 | 3.771 | 3.439 | 4.136 | 3.757 | 3.415 | 4.132 | 2.204 | 1.938 | 2.506 | 2.071 | 1.797 | 2.386 | 9074.241 | 0.174 |
| PASC: At Least 3 Positive | 2257/21.9 | 3.770 | 3.417 | 4.160 | 3.757 | 3.395 | 4.157 | 2.040 | 1.756 | 2.370 | 2.059 | 1.769 | 2.395 | 7940.332 | 0.158 |

Model 1: adjusted for age, sex and race
Model 2: Model 1 + bmi, comorbidities, boosted COVID-19 vaccination status, and boosted COVID-19 vaccination status/OSA interaction
Model 3: Model 2 + income, employment and education

*See text for definition of PASC Models
