## Supplementary material for "Association of Obstructive Sleep Apnea with Post-Acute Sequelae of SARS-CoV-2 infection (PASC)": Table S1

**Table S1: Logistic Regression Models of the Association of Obstructive Sleep Apnea with Overall PASC Models, Sensitivity Analyses**

| | Unadjusted | | | Fully Adjusted* | | |
|---|---|---|---|---|---|---|
| | *OR* | *95% CI* | | *aOR* | *95% CI* | |
| **PASC Model** | | *Lower* | *Upper* | | *Lower* | *Upper* |
| *COVID-19 Infection=Positive Test or Loss of Taste or Smell (N=9853)* | | | | | | |
| PASC: 95th Percentile | 3.688 | 3.364 | 4.043 | 2.020 | 1.755 | 2.326 |
| PASC: 99th Percentile | 3.350 | 3.033 | 3.699 | 1.983 | 1.665 | 2.256 |
| PASC: At Least 2 Positive | 3.672 | 3.344 | 4.032 | 2.058 | 1.784 | 2.375 |
| PASC: At Least 3 Positive | 3.647 | 3.301 | 4.028 | 2.040 | 1.751 | 2.376 |
| *COVID-19 Infection=Positive Test, Loss of Taste or Smell, Clinical Diagnosis (No Positive Test), Self-reported (No Positive Test) (N=11521)* | | | | | | |
| PASC: 95th Percentile | 3.860 | 3.537 | 4.212 | 2.014 | 1.762 | 2.303 |
| PASC: 99th Percentile | 3.615 | 3.284 | 3.979 | 1.958 | 1.689 | 2.269 |
| PASC: At Least 2 Positive | 3.937 | 3.599 | 4.307 | 2.105 | 1.834 | 2.416 |
| PASC: At Least 3 Positive | 3.947 | 3.585 | 4.345 | 2.110 | 1.819 | 2.447 |
| *Obstructive Sleep Apnea=Current or Past Treatment (n=10324)†* | | | | | | |
| PASC: 95th Percentile | 2.209 | 1.924 | 2.536 | 1.120 | 0.923 | 1.358 |
| PASC: 99th Percentile | 2.148 | 1.858 | 2.483 | 1.229 | 0.984 | 1.535 |
| PASC: At Least 2 Positive | 2.323 | 2.022 | 2.668 | 1.268 | 1.047 | 1.534 |
| PASC: At Least 3 Positive | 2.285 | 0.001 | 2.640 | 1.277 | 1.022 | 1.597 |
| *Obstructive Sleep Apnea=Treated (N=10324)†* | | | | | | |
| PASC: 95th Percentile | 1.136 | 0.974 | 1.325 | 1.106 | 0.881 | 1.389 |
| PASC: 99th Percentile | 0.976 | 0.832 | 1.145 | 0.983 | 0.784 | 1.232 |
| PASC: At Least 2 Positive | 0.986 | 0.846 | 1.149 | 0.927 | 0.743 | 1.157 |
| PASC: At Least 3 Positive | 0.941 | 0.803 | 1.103 | 0.934 | 0.744 | 2.396 |

*adjusted for age, sex, race, bmi, comorbidities, boosted COVID-19 vaccination status, and boosted COVID-19 vaccination status/OSA interaction, income, employment and education

†COVID-19 Infection=Positive test, loss of taste or smell, or received a diagnosis without testing positive
